# Prevalence of Financial Toxicity and Its Association with Medication Adherence Among Patients Undergoing Haemodialysis Under Comprehensive Public Support in Japan

**DOI:** 10.1101/2025.07.23.25332060

**Authors:** Ryohei Inanaga, Tatsunori Toida, Tetsuro Aita, Yusuke Kanakubo, Mamiko Ukai, Takumi Toishi, Atsuro Kawaji, Masatoshi Matsunami, Tadao Okada, Yu Munakata, Tomo Suzuki, Noriaki Kurita

## Abstract

**Background and hypothesis:** Financial toxicity (FT) refers not only to the difficulty in affording medical care but also to the psychological distress and perceived financial burden it imposes. Although dialysis in Japan is extensively covered by public insurance, little is known about the prevalence of FT and its effects on medication adherence. This study aimed to assess the prevalence of FT and examine its association with medication adherence among patients undergoing haemodialysis in Japan.

**Methods:** This multicentre, cross-sectional study included Japanese adults undergoing in-centre haemodialysis at six facilities. FT was assessed using the Comprehensive Score for Financial Toxicity (COST), and medication adherence was assessed using the 12-item Adherence Starts with Knowledge (ASK-12) scale. The COST scores were compared with published data from Japanese patients with cancer and patients undergoing dialysis from other countries using unpaired t-tests. Associations between the COST and ASK-12 scores were analysed using multivariate general linear models.

**Results:** In total, 455 participants were included in the analysis. The mean COST score was 22.0, and 68% of the participants experienced at least mild FT. FT severity was comparable to that of Japanese patients with cancer and significantly lower than that reported among patients undergoing dialysis in Brazil and China. Lower FT (i.e., higher COST scores) was associated with fewer medication adherence difficulties (per 1-point higher: β = –0.19). This association was particularly evident in the ‘inconvenience/forgetfulness’ and ‘behaviour’ subdomains (per 1-point higher: β = –0.06 and β = –0.08, respectively).

**Conclusions:** Despite generous public coverage, FT is common among Japanese patients undergoing haemodialysis and is associated with difficulties in medication adherence. The awareness of hidden financial distress and its integration into shared decision-making regarding prescriptions may help improve treatment adherence and patient outcomes.

**Key learning points:** *What was known:*

- Among patients undergoing dialysis, medication adherence rates are generally < 70%.
- ‘Financial toxicity’ is associated with poor medication adherence in oncology.
- However, among patients undergoing dialysis receiving publicly funded care, the prevalence of financial toxicity and its effects on medication adherence remain unclear.

*This study adds:*

- Despite Japan’s comprehensive public insurance system, approximately 70% of patients undergoing dialysis experience at least mild financial toxicity levels, comparable to those observed in Japanese patients with cancer.
- Lower financial toxicity was associated with better medication adherence, particularly in the ‘inconvenience/forgetfulness’ and ‘behaviour’ subdomains.

*Potential impact:*

- Financial hardship can cause stress. Healthcare providers should build trust with their patients and foster open discussions on financial and social challenges.
- Practising shared decision-making is essential for prescribing medications that consider patients’ financial burden.
- Providing work-friendly dialysis schedules may support patients’ long-term financial independence.

## Introduction

Poor medication adherence in patients with chronic kidney disease (CKD) is associated with higher mortality and hospitalisation rates [1, 2], making its improvement a critical clinical goal. Among patients undergoing dialysis, medication adherence is often suboptimal—typically < 70%—owing to the complexity of treatment regimens requiring > 10 medications daily [3–5]. However, medication adherence issues are not solely attributable to the pill burden; financial hardship also plays a significant role [6].

High out-of-pocket costs and difficulties in affording medications contribute to poor adherence among patients undergoing dialysis and patients with CKD [7–9]. This burden extends beyond affordability, encompassing psychological stress and a sense of financial distress, collectively termed ‘financial toxicity’ (FT) [10]. In oncology, FT is commonly assessed using the Comprehensive Score for Financial Toxicity (COST) [11] and is associated with treatment discontinuation [12, 13]. Despite its relevance, little is known about the prevalence of FT in patients undergoing dialysis or its effects on medication adherence in this population. Studies from Brazil and China have reported a high prevalence of FT among patients undergoing haemodialysis (HD) and peritoneal dialysis (PD), respectively [14, 15]. Although these countries offer publicly funded dialysis under their national health systems, disparities in regional coverage and relatively lower national incomes may contribute to increased FT in these populations. Even in high-income countries, such as the United States, many patients with end-stage kidney disease (ESKD) experience FT [16]. However, these studies did not examine the effects of FT on medication adherence.

In contrast, Japan provides universally accessible, publicly funded dialysis, with out-of-pocket costs typically capped at approximately 10,000 yen per month. Despite this generous coverage, patients may experience anxiety and psychological distress owing to concerns about future income loss and financial burden on their families [17]. This suggests that even in a high-income country with comprehensive medical support, the subjective burden of FT may persist and potentially affect health behaviours. Understanding how financial distress, beyond affordability, affects medication adherence among patients undergoing dialysis in Japan is essential. Such insights can inform the development of targeted support systems and guide the training of healthcare providers to address patients’ financial concerns better.

Against this background, the present multicentre cross-sectional study aimed to (1) assess the prevalence of FT using the COST and (2) examine its association with medication adherence among patients undergoing HD in Japan.

## Materials and Methods

### Study design and participants

This multicentre cross-sectional study was conducted at six medical facilities providing outpatient maintenance dialysis. Participants were eligible if they (1) were adults with ESKD undergoing HD or a combination of HD and PD, (2) received dialysis treatment at one of the participating facilities, or (3) were able to complete a self-administered questionnaire. Written informed consent was obtained from all the participants. The participants completed a paper-based questionnaire, which was collected in a manner that prevented healthcare professionals from viewing the responses. Respondents were provided with a gift card worth 500 yen. The data were collected between April 2022 and February 2023. This study followed the Declaration of Helsinki and was approved by the Ethical Review Board of Fukushima Medical University (approval number: ippan2021-292).

## Measures

### Outcome: Medication adherence

The primary outcome was medication adherence, which was assessed using the Japanese version of the Adherence Starts with Knowledge (ASK)-12 scale [18, 19]. The ASK-12 is a 12-item scale categorised into three subdomains: (inconvenience/forgetfulness [three items], belief in treatment [four items], and behaviour [five items]) (Box S1). Each item is scored on a 5-point Likert scale, with scores ranging from 1 (‘very often’) to 5 (‘never’). Items 4–7 were reverse-scored. The total score ranges from 12 to 60 points, with the following subdomain scores: inconvenience/forgetfulness (3–15 points), belief (4–20 points), and behaviour (5–25 points). Higher scores indicate greater adherence difficulties. The reliability (Cronbach’s α = 0.75) and validity of the ASK-12 scale have been confirmed [18], and the Japanese version of the ASK-12 scale has demonstrated criterion-related validity in studies of patients with asthma through associations with refill rates at pharmacies and attack frequency [19, 20].

### Exposure: Financial toxicity

FT, the main exposure, was assessed using the Japanese version of the COST [11] (Box S2). The COST is an 11-item self-report questionnaire, with each item scored on a 5-point Likert scale (0 = ‘not at all’ to 4 = ‘very much’). Items 2–5 and 8-10 were reverse-scored. The total scores range from 0 to 44, with lower scores indicating greater FT. The COST score is categorised into four grades of FT: G0 (non-toxic, ≥ 26), G1 (mild, 14–25), G2 (moderate, 1–13), and G3 (severe, 0). Based on this classification, patients with a grade of G1 or higher were defined as experiencing FT [21]. The original scale demonstrated strong construct validity and internal consistency (Cronbach’s α = 0.92) [22]. The Japanese version also demonstrated internal consistency (α = 0.87) and construct validity in studies of patients with cancer [23, 24].

### Measurement of covariates

Covariates were selected based on the literature and expert medical knowledge, focusing on variables that may influence financial toxicity and medication adherence. The following covariates were included: age, sex, smoking history, final education, household income, comorbidities (hypertension, diabetes, cardiovascular disease, liver disease, depression, dementia), dialysis time per session, number of dialysis sessions per week, use of PD, total number of classes of antihypertensive medications, and number of phosphate binders prescribed. Cardiovascular disease was defined as a history of myocardial infarction, angina pectoris, heart failure, cerebrovascular disease, or peripheral vascular diseases. Antihypertensive medications were classified into the following five classes, and the presence of any one class was included: (i) calcium channel blockers; (ii) angiotensin-converting enzyme inhibitors, angiotensin II receptor blockers, or angiotensin receptor neprilysin inhibitors, (iii) alpha-beta or beta blockers; (iv) alpha blockers; and (v) central sympathetic nerve blockers. The pill count for the phosphate binders was calculated if any of the following were prescribed: calcium carbonate, sevelamer, bixalomer, lanthanum, sucroferric oxyhydroxide, or ferrous citrate hydrate.

### Statistical analyses

Patient characteristics are described as medians and interquartile ranges (IQRs) for continuous variables and as frequencies and proportions for categorical variables. We compared the COST scores of our HD cohort with those of Japanese patients with cancer [23], Brazilian patients undergoing HD [14], and Chinese patients undergoing PD [15] using unpaired t-tests based on previously reported means and standard deviations. To examine the association between FT and medication adherence, we performed general linear regression analyses adjusted for the covariates listed above and analysed both the total and subdomain scores for medication adherence. Missing data were assumed to be missing at random and handled using multiple imputations with chained equations (20 imputed datasets). Statistical significance was set at P = 0.05. All analyses were performed using Stata/SE version 17 (StataCorp, College Station, TX, USA).

## Results

### Study flow and participant characteristics

Of the 651 patients undergoing HD across the six facilities, 484 (74.3%) consented to participate. After excluding 28 patients with missing outcome data and one patient without a regular prescription, 455 patients were included in the final analysis (Figure 1). Table 1 summarises these characteristics. The median age was 71 (IQR, 62–78) years, and 300 (65.9%) participants were male. More than half (53.8%) of the participants reported an annual household income of < 3 million yen. The median dialysis vintage was 5.7 (IQR, 2.9–10.6) years, and 3.5% were also undergoing PD.

**Figure 1.**
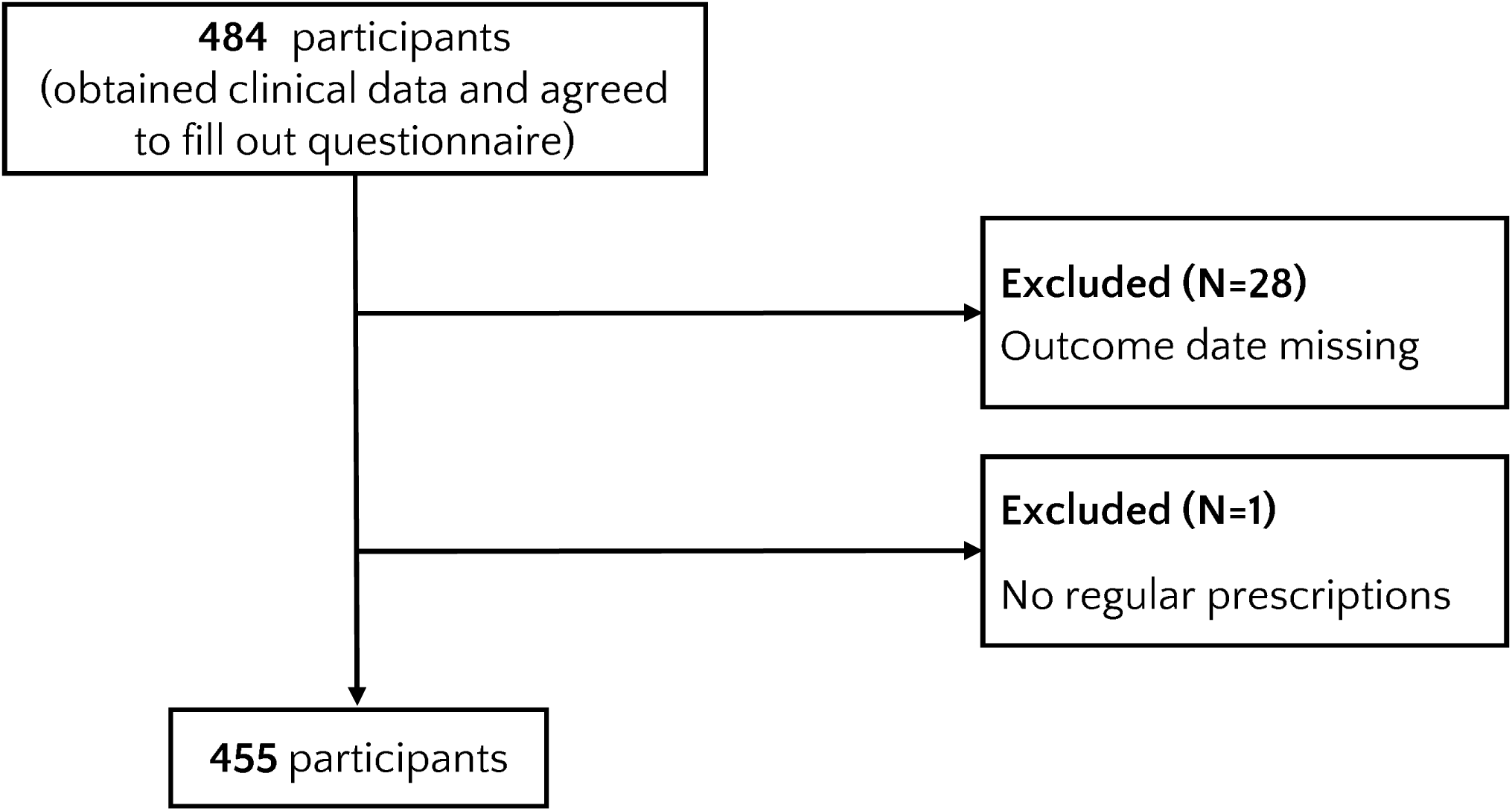
Flowchart of study design.

**Table 1.** Patient characteristics (n = 455)

|  | **Total n = 455** | **Missing,**<br>**n** |
| --- | --- | --- |
| Age (years) | 71 [62–78] | 0 |
| Men, n (%) | 300 (65.9) | 0 |
| Dialysis vintage (years) | 5.7 [2.9–10.6] | 0 |
| Renal disease, n (%) |  | 0 |
| *Diabetic nephropathy* | 171 (37.6) |  |
| *Chronic glomerulonephritis* | 106 (23.3) |  |
| *Nephrosclerosis* | 60 (13.2) |  |
| *Polycystic kidney disease* | 13 (2.9) |  |
| *Others* | 69 (15.1) |  |
| *Unknown* | 36 (7.9) |  |
| Comorbidities, n (%) |  |  |
| *Hypertension* | 349 (76.7) | 0 |
| *Diabetes* | 195 (43.1) | 3 |
| *Liver disease* | 27 (6.0) | 1 |
| *Cardiovascular disease* | 195 (43.2) | 4 |
| *Depression* | 10 (2.2) | 1 |
| *Dementia* | 20 (4.4) | 3 |
| Dialysis time per session, n (%) |  | 6 |
| < 4.0 h | 55 (12.2) |  |
| 4.0–< 5.0 h | 381 (84.9) |  |
| ≥ 5.0 h | 13 (2.9) |  |
| Dialysis frequency per week, n (%) |  | 2 |
| *Once* | 14 (3.1) |  |
| *Twice* | 8 (1.8) |  |
| *Three times* | 430 (94.9) |  |
| *Four times* | 1 (0.2) |  |
| Combined therapy with peritoneal dialysis, n (%) | 16 (3.5) | 0 |
| Smoking, n (%) |  | 19 |
| *Never* | 233 (53.4) |  |
| *Ever* | 153 (35.1) |  |
| *Current* | 50 (11.5) |  |
| Education, n (%) |  | 20 |
| *Junior high school graduate or less* | 105 (24.2) |  |
| *High school graduate* | 212 (48.7) |  |
| *University/graduate school graduate* | 57 (14.0) |  |
| *Others* | 61 (13.1) |  |
| Household income, n (%) |  | 33 |
| < 1,000,000 yen | 41 (9.7) |  |
| 1,000,000–< 3,000,000 yen | 186 (44.1) |  |
| 3,000,000–< 5,000,000 yen | 111 (26.3) |  |
| 5,000,000–< 10,000,000 yen | 70 (16.6) |  |
| $\geq 10,000,000$ yen | 14 (3.3) | |
| Number of phosphorus binder, n (%) |  | 1 |
| *None* | 62 (13.7) |  |
| *1–3 tablets* | 135 (29.7) |  |
| *$\geq 4$ tablets* | 257 (56.6) |  |
| Number of antihypertensive classes, n (%) |  | 0 |
| *None* | 71 (15.6) |  |
| *One* | 126 (27.7) |  |
| *Two* | 171 (37.6) |  |
| *Three or more* | 87 (19.1) |  |

- 1 Continuous variables are summarised as the medians [interquartile ranges]. - 2 Categorical variables are summarised as frequencies and proportions (in parentheses). - 3

### Comparison of financial toxicity: Japanese patients with Cancer and patients undergoing dialysis in other countries

The mean COST score was 22.0 (standard deviation, 6.8), and 68% experienced at least mild FT. This level of FT did not significantly differ from that of Japanese patients with cancer (P = 0.23; mean difference, 0.9; 95% confidence interval [CI], –0.57 to 2.37; effect size [ES], 0.12) [23]. In contrast, the COST score was significantly higher than that reported for patients undergoing HD in Brazil (P = 0.01; mean difference, 1.7; 95% CI, 0.37 to 3.03) [14] and patients undergoing PD in China (P < 0.01; mean difference, 6.7; 95% CI, 5.58 to 7.92) [15] (Figure 2). The magnitude of the difference was small compared with that in Brazilian patients undergoing HD (ES = 0.23) and larger than that in Chinese patients undergoing PD (ES = 1.02).

**Figure 2.**
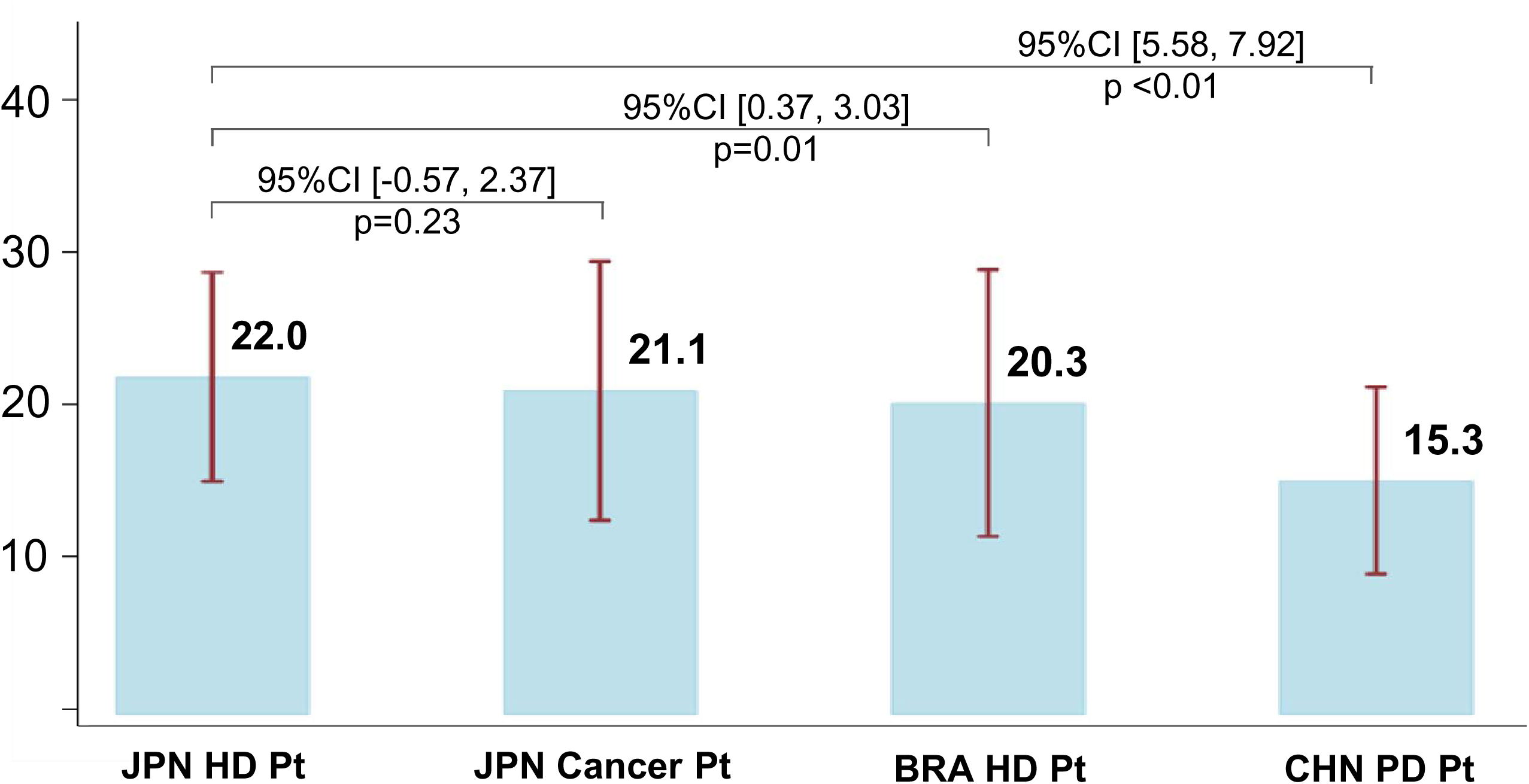
Comparison of mean Comprehensive Score for Financial Toxicity (COST) scores among patient groups. Japanese patients undergoing haemodialysis had higher COST scores than Brazilian patients undergoing haemodialysis and Chinese patients undergoing peritoneal dialysis but similar scores to Japanese patients with cancer. Higher scores indicated lower financial toxicity. Error bars represent the standard deviation.

### Association between financial toxicity and medication adherence

The median total ASK-12 score was 23 (IQR, 19–28). The median scores for each domain were as follows: inconvenience/forgetfulness, 6 (IQR, 4–8); health beliefs, 10 (IQR, 8–12); and behaviour, 6 (IQR, 5–9). Table 2 presents the associations between the total ASK-12 scores, COST scores, and covariates. Lower FT (higher COST score) was associated with better medication adherence; each 1-point increase in COST score corresponded to a 0.19 point decrease in adherence difficulty (β = –0.19; 95% CI, –0.29 to –0.09). Longer dialysis sessions (≥ 5 h) were associated with greater difficulty compared with sessions < 4 h (β = 4.01; 95% CI, 0.04 to 7.99). Patients taking only one antihypertensive medication also reported more difficulty than those taking none (β = 2.01; 95% CI, 0.17 to 3.84). Table 3 lists domain-specific associations.

**Table 2.** Association between medication adherence and financial toxicity and other covariates^a^ (n = 455)

|  | Mean difference, point estimate<br>(95% CI) | P-value |
| --- | --- | --- |
| Financial toxicity, per 1-pt increase | **−0.19 (−0.29 to −0.09)** | **< 0.01** |
| Age, per 10-yr increase | 0.08 (−0.45 to 0.61) | 0.77 |
| Women vs. men | −1.04 (−2.48 to 0.40) | 0.16 |
| Comorbidities |  |  |
| *Hypertension* | 0.61 (−0.78 to 2.00) | 0.39 |
| *Diabetes* | −0.14 (−1.36 to 1.08) | 0.82 |
| *Liver disease* | 0.58 (−1.85 to 3.00) | 0.64 |
| *Cerebrovascular disease* | 0.21 (−1.00 to 1.43) | 0.73 |
| *Depression* | 0.25 (−3.71 to 4.21) | 0.90 |
| *Dementia* | 0.31 (−2.52 to 3.13) | 0.83 |
| Dialysis time per session, |  |  |
| < 4.0 h | Reference |  |
| 4.0–< 5.0 h | 0.6 (−1.30 to 2.50) | 0.54 |
| ≥ 5.0 h | **4.01 (0.04 to 7.99)** | **0.048** |
| Dialysis frequency per week |  |  |
| *Once* | Reference |  |
| *Twice* | −1.34 (−11.57 to 8.89) | 0.80 |
| *Three times* | 0.6 (−8.57 to 9.77) | 0.90 |
| *Four times* | −3.53 (−18.89 to 11.83) | 0.65 |
| Combined therapy with peritoneal dialysis | 0.33 (−8.23 to 8.90) | 0.94 |
| Smoking |  |  |
| *Never* | Reference |  |
| *Ever* | −0.33 (−1.79 to 1.13) | 0.66 |
| *Current* | 0.41 (−1.67 to 2.49) | 0.70 |
| Number of antihypertensive classes |  |  |
| *None* | Reference |  |
| *One* | **2.01 (0.17 to 3.84)** | **0.03** |
| *Two* | 1.57 (−0.19 to 3.33) | 0.08 |
| *Three or more* | 0.14 (−1.88 to 2.17) | 0.89 |
| Education |  |  |
| *Junior high school graduate or less* | Reference |  |
| *High school graduate* | 1.29 (−0.30 to 2.88) | 0.11 |
| *University/graduate school graduate* | 1.44 (−0.80 to 3.67) | 0.21 |
| *Others* | 1.12 (−0.96 to 3.20) | 0.29 |
| Household Income |  |  |
| < 1,000,000 yen | Reference |  |
| 1,000,000–< 3,000,000 yen | −0.06 (−2.21 to 2.10) | 0.96 |
| 3,000,000–< 5,000,000 yen | −1.01 (−3.32 to 1.29) | 0.39 |
| 5,000,000–< 10,000,000 yen | 0.2 (−2.37 to 2.76) | 0.88 |
| *≥ 10,000,000 yen* | 3.47 (−0.58 to 7.53) | 0.09 |
| Phosphate binder |  |  |
| *None* | Reference |  |
| *1–3 tablets* | −0.14 (−2.04 to 1.76) | 0.89 |
| *≥ 4 tablets* | 0.46 (−1.33 to 2.25) | 0.61 |

1 Values in bold are statistically significant.
<sup>a</sup>A general linear model was fitted with the inclusion of all variables listed above.
CI, confidence interval

**Table 3.** Association between medication adherence subdomain and financial toxicity and other covariates^a^ (n = 455)

|  | Subdomain 1 = inconvenience |  | Subdomain 2 = belief |  | Subdomain 3 = behaviour |  |
| --- | --- | --- | --- | --- | --- | --- |
|  | Mean difference, point estimate (95% CI) | P-value | Mean difference, point estimate (95% CI) | P-value | Mean difference, point estimate (95% CI) | P-value |
| Financial toxicity, per 1-pt increase | **−0.06 (−0.11 to −0.02)** | **< 0.01** | −0.04 (−0.09 to 0.003) | 0.07 | **−0.08 (−0.14 to −0.03)** | **< 0.01** |
| Age, per 10-yr increase | 0.1 (−0.13 to 0.33) | 0.39 | 0.01 (−0.25 to 0.26) | 0.96 | −0.03 (−0.33 to 0.27) | 0.85 |
| Women vs. men | −0.45 (−1.07 to 0.18) | 0.16 | −0.01 (−0.69 to 0.67) | 0.98 | −0.59 (−1.4 to 0.23) | 0.16 |
| Comorbidities |  |  |  |  |  |  |
| *Hypertension* | 0.13 (−0.47 to 0.73) | 0.67 | −0.04 (−0.7 to 0.61) | 0.90 | 0.52 (−0.27 to 1.31) | 0.20 |
| *Diabetes* | −0.07 (−0.6 to 0.46) | 0.80 | 0.05 (−0.52 to 0.62) | 0.86 | −0.12 (−0.82 to 0.57) | 0.73 |
| *Liver disease* | 0.61 (−0.44 to 1.66) | 0.25 | 0.17 (−0.97 to 1.31) | 0.77 | −0.21 (−1.59 to 1.17) | 0.77 |
| *Cerebrovascular disease* | 0.09 (−0.44 to 0.61) | 0.75 | 0.29 (−0.28 to 0.87) | 0.31 | −0.17 (−0.86 to 0.52) | 0.63 |
| *Depression* | −0.4 (−2.12 to 1.32) | 0.65 | 1.25 (−0.62 to 3.12) | 0.19 | −0.6 (−2.86 to 1.66) | 0.60 |
| *Dementia* | 1.19 (−0.03 to 2.41) | 0.06 | −0.47 (−1.81 to 0.87) | 0.49 | −0.41 (−2.01 to 1.19) | 0.62 |
| Dialysis time per session, |  |  |  |  |  |  |
| < 4.0 h | Reference |  | Reference |  | Reference |  |
| 4.0–< 5.0 h | 0.52 (−0.31 to 1.35) | 0.22 | 0.35 (−0.55 to 1.24) | 0.45 | −0.27 (−1.34 to 0.81) | 0.63 |
| ≥ 5.0 h | 1.16 (−0.57 to 2.89) | 0.19 | **2.18 (0.3 to 4.06)** | **0.02** | 0.67 (−1.59 to 2.93) | 0.56 |
| Dialysis frequency per week |  |  |  |  |  |  |
| *Once* | Reference |  | Reference |  | Reference |  |
| *Twice* | −0.27 (−4.71 to 4.17) | 0.91 | 3.21 (−1.61 to 8.03) | 0.19 | −4.28 (−10.09 to 1.53) | 0.15 |
| *Three times* | 0.22 (−3.76 to 4.2) | 0.91 | 2.91 (−1.42 to 7.23) | 0.19 | −2.53 (−7.74 to 2.69) | 0.34 |
| *Four times* | 0.84 (−5.82 to 7.51) | 0.80 | 0.81 (−6.43 to 8.05) | 0.83 | −5.18 (−13.91 to 3.55) | 0.24 |
| Combined therapy with peritoneal dialysis | 0.37 (−3.35 to 4.08) | 0.84 | 2.74 (−1.3 to 6.77) | 0.18 | −2.77 (−7.64 to 2.1) | 0.26 |
| Smoking |  |  |  |  |  |  |
| *Never* | Reference |  | Reference |  | Reference |  |
| *Ever* | -0.1 (-0.72 to 0.53) | 0.76 | -0.28 (-0.97 to 0.4) | 0.42 | 0.05 (-0.77 to 0.87) | 0.91 |
| *Current* | -0.08 (-0.98 to 0.82) | 0.86 | 0.03 (-0.95 to 1) | 0.96 | 0.46 (-0.69 to 1.62) | 0.43 |
| Number of antihypertensive classes |  |  |  |  |  |  |
| *None* | Reference |  | Reference |  | Reference |  |
| *One* | **0.95 (0.15 to 1.74)** | **0.02** | 0.59 (-0.28 to 1.45) | 0.18 | 0.47 (-0.57 to 1.51) | 0.37 |
| *Two* | **0.95 (0.18 to 1.71)** | **0.02** | 0.23 (-0.60 to 1.06) | 0.59 | 0.4 (-0.60 to 1.39) | 0.44 |
| *Three or more* | 0.41 (-0.47 to 1.29) | 0.36 | 0.06 (-0.89 to 1.02) | 0.90 | -0.32 (-1.48 to 0.83) | 0.58 |
| Education |  |  |  |  |  |  |
| *Junior high school graduate or less* | Reference |  | Reference |  | Reference |  |
| *High school graduate* | 0.12 (-0.59 to 0.83) | 0.74 | 0.53 (-0.22 to 1.28) | 0.16 | 0.64 (-0.28 to 1.56) | 0.18 |
| *University/graduate school graduate* | 0.03 (-0.93 to 0.99) | 0.95 | 0.17 (-0.84 to 1.18) | 0.74 | 1.24 (-0.07 to 2.55) | 0.06 |
| *Others* | 0.11 (-0.80 to 1.02) | 0.81 | -0.07 (-1.03 to 0.88) | 0.88 | 1.08 (-0.11 to 2.28) | 0.08 |
| Household Income |  |  |  |  |  |  |
| *< 1,000,000 yen* | Reference |  | Reference |  | Reference |  |
| *1,000,000–< 3,000,000 yen* | -0.1 (-1.04 to 0.83) | 0.83 | -0.68 (-1.68 to 0.32) | 0.18 | 0.72 (-0.53 to 1.97) | 0.26 |
| *3,000,000–< 5,000,000 yen* | -0.45 (-1.45 to 0.54) | 0.37 | -0.68 (-1.78 to 0.42) | 0.23 | 0.12 (-1.22 to 1.45) | 0.86 |
| *5,000,000–< 10,000,000 yen* | -0.02 (-1.16 to 1.11) | 0.97 | -0.58 (-1.77 to 0.60) | 0.34 | 0.8 (-0.68 to 2.29) | 0.29 |
| *≥ 10,000,000 yen* | 1.35 (-0.41 to 3.10) | 0.13 | -0.35 (-2.17 to 1.48) | 0.71 | **2.47 (0.17 to 4.77)** | **0.04** |
| Phosphate binder |  |  |  |  |  |  |
| *None* | Reference |  | Reference |  | Reference |  |
| *1 to 3 tablets* | -0.13 (-0.95 to 0.69) | 0.75 | 0.02 (-0.87 to 0.91) | 0.97 | -0.03 (-1.10 to 1.05) | 0.96 |
| *4 or more tablets* | 0.18 (-0.59 to 0.96) | 0.65 | -0.39 (-1.23 to 0.45) | 0.36 | 0.67 (-0.34 to 1.69) | 0.19 |

1 Values in bold are statistically significant.
<sup>a</sup>For each subdomain score, a general linear model was fitted with the inclusion of all variables listed above.
CI, confidence interval

Higher COST scores were associated with lower difficulty in the inconvenience domain (β per point = –0.06; 95% CI, –0.11 to –0.02) and behaviour domain (β = –0.08; 95% CI, –0.14 to – 0.03), suggesting that lower FT is associated with fewer barriers in these specific aspects of adherence.

## Discussion

This study demonstrated that even in Japan, where dialysis is universally covered by public funding, many patients undergoing HD experience FT with severity comparable to that of patients with cancer. Furthermore, FT was associated with greater difficulty in medication adherence, particularly in the ‘inconvenience/forgetfulness’ and ‘behaviour’ subdomains.

Our findings support and extend the previous research in several ways. First, we showed that Japanese patients undergoing dialysis experienced a level of FT comparable to patients with cancer in Japan. This suggests that ESKD imposes not only a survival burden but also a substantial financial burden. In Japan, out-of-pocket expenses for dialysis are capped at ¥10,000–¥20,000 per month, whereas patients with cancer often incur higher direct medical costs. Despite this, the comparable FT observed may reflect the broader socioeconomic burden of dialysis, including treatment-related work limitations, reduced social roles, and dissatisfaction with income owing to the inability to work, rather than direct costs alone.

In contrast, patients on dialysis in Brazil and China reported greater FT than those in our cohort, as measured using the COST instrument. This discrepancy may be attributed to differences in national income levels and socioeconomic status. For example, in a Brazilian study, approximately one-quarter of the participants had household incomes of just one to two times the minimum wage [14]. In a Chinese study [15], the cohort was relatively young (mean age, 48 years) but had low employment rates, and nearly 30% were uninsured or enrolled in rural insurance plans with higher out-of-pocket costs [25]. Similarly, high levels of FT have been reported among patients with ESKD in the United States, a high-income country [16]. This may be attributed to the substantial out-of-pocket costs under Medicare coverage or the inclusion of hospitalised patients who may have been experiencing acute financial stress at the time of the assessment.

Second, our finding that higher FT was associated with greater difficulty in medication adherence, particularly forgetfulness, adds a novel dimension to the existing literature. Previous studies, such as those among US Medicare beneficiaries or multinational comparisons across 12 countries, have focused primarily on direct medication costs and behavioural adherence indicators (e.g. skipped doses or unfilled prescriptions) [7, 8]. Although qualitative studies have hypothesised that financial strain can lead to self-adjusted medication behaviours or discontinuation, our study quantitatively demonstrated that FT might impair adherence through cognitive or psychological mechanisms, even when medications are affordable [9].

This study has several important implications for clinicians and policymakers. First, the observed association between FT and medication non-adherence, particularly forgetfulness and inconvenience, suggests that missed doses among patients undergoing dialysis may not simply reflect inattentiveness or a lack of motivation. Rather, such behaviour may be shaped by the patients’ socioeconomic environments. This aligns with the cognitive burden of poverty theory, which posits that sustained financial hardship induces chronic stress, impairing executive function and self-care behaviours [26]. Thus, clinicians should foster trusting relationships that enable open discussions regarding financial difficulties and other social challenges. Second, the association between FT and difficulties in medication-related behaviour underscores the need for a regular review of prescriptions considering treatment affordability. Engaging patients in shared decision-making regarding medication costs may facilitate safe regimen simplification or selection of more affordable alternatives without compromising disease control. Third, educational programmes aimed at improving patients’ financial literacy and budgeting skills should be considered to promote long-term financial resilience. Such support—through household expense management and counselling—may reduce economic stress and improve the psychological capacity for continued medication adherence [17, 27]. Finally, for working patients undergoing dialysis, flexible dialysis scheduling aligned with employment demands may help sustain workforce participation. Such accommodations are effective when dialysis providers collaborate closely with employers [28].

This study has several strengths. First, the inclusion of multiple dialysis centres across urban and suburban regions, combined with a high participation rate and sufficient sample size, enhances the generalisability of the findings to the national dialysis population. Second, despite Japan’s extensive public financial support for dialysis care, this study revealed a substantial prevalence of FT, highlighting not only economic burdens but also the associated social and psychological distress. Third, using the ASK-12 scale and analysing its three distinct domains—behaviour, barriers, and inconvenience/forgetfulness—this study provides granular insights into the specific aspects of medication adherence that are most affected by FT.

This study has some limitations. First, its cross-sectional design precluded causal inferences. Second, medication adherence was assessed by self-reporting and did not account for factors such as pill burden, regimen complexity, or formulation type. Although more objective methods, such as the Medication Event Monitoring System, exist, their application in large-scale epidemiological studies is often impractical because of cost and training demands [29]. Third, the findings were derived from Japan, where public health insurance minimises out-of-pocket costs; thus, generalisability to countries with higher cost sharing may be limited. However, studies conducted in such settings are likely to observe even more pronounced FT and medication adherence difficulties [8]. Finally, data on employment history and insurance status were unavailable. Although these factors may influence FT, their direct effect on medication adherence remains unclear.

In conclusion, despite the extensive public financial support available to patients undergoing HD in Japan, FT remains prevalent and is significantly associated with difficulties in medication adherence, particularly within the domains of ‘inconvenience/forgetfulness’ and ‘behaviour’. These findings underscore the need for dialysis providers to consider the potential influence of unspoken financial distress when addressing adherence. Rather than attributing non-adherence solely to a lack of patient motivation, clinicians should strive to create a supportive clinical environment in which patients feel safe disclosing their financial concerns. Such efforts may contribute to more patient-centred decision-making regarding medication management.

## Supporting information

Supplementary Box S1, S2

## Data Availability

The datasets generated and/or analysed in the current study are available from the corresponding author upon reasonable request.

## Acknowledgements

The authors greatly thank the following researchers, research assistants, and medical staff members for their assistance in collecting the questionnaire-based and clinical information used in this study: Ms. Aki Tairaku (Shin-Yurigaoka General Hospital,Kawasaki-City, Kanagawa); Ms. Takako Saruwatari and Ms. Akiko Kamimura (Kyushu University of Health and Welfare, Nobeoka-City, Miyazaki); Tetuo Ueki, MD, Akio Munakata, MD, Yoshihiko Watanabe, MD (Munakata Clinic, Mobara-City, Chiba); Ms.Yayoi Takanashi, Reiji Masaki, NP, Tomohiko Inoue, MD, Shinnosuke Sugihara, MD, Kanako Nagaoka, MD and Hiroshi Kuji, MD (Kameda Medical Center, Kamogawa-City,Chiba); Kenji Yamaguchi, MD (Awa Regional Medical Center, Tateyama-City, Chiba); Ms. Miyuki Sato (Fukushima Medical University Hospital, Fukushima-City, Fukushima).

## Authors’ Contributions

Research idea and study design: RI, TT, and NK; data acquisition: RI, YK, MU, MM, TO, YM, and TS; data analysis/interpretation: TA and NK; statistical analyses: RI, TT, and NK; and supervision or mentorship: TT and NK. Each author contributed important intellectual content during manuscript drafting or revision, agreed to be personally accountable for the individual’s contributions, and ensured that questions on the accuracy or integrity of any portion of the work, even one in which the author was not directly involved, were appropriately investigated and resolved, including documentation in the literature, if appropriate.

## Supplementary Materials

**Box S1**. Japanese version of the Adherence Starts with Knowledge 12 (ASK-12) scale

**Box S2**. Japanese version of the Comprehensives Score for Financial Toxicity (COST)

