## Supplementary Box S1, S2 for "Prevalence of Financial Toxicity and Its Association with Medication Adherence Among Patients Undergoing Haemodialysis Under Comprehensive Public Support in Japan"

**Supplementary Materials**

**Box S1. Japanese version of the Adherence Starts with Knowledge 12 (ASK-12) scale**

**Box S2. Japanese version of the Comprehensive Score for Financial Toxicity (COST)**

**Box S1. Japanese version of the Adherence Starts with Knowledge 12 (ASK-12) scale** ^1,2^

| Instruction sentence | Taking Medicine-What Gets in the Way? Think about all of the medicines you take. Mark one answer for each item below. |
| --- | --- |
| Lifestyles | |
| Question 1 | I forget to take my medicines some of the time. |
| Question 2 | I run out of my medicines because I don’t get refills on time. |
| Question 3 | Taking medicines more than once a day is inconvenient. |
| Attitudes and Beliefs | |
| Question 4 | I feel confident that each one of my medicines will help  me. |
| Question 5 | I know if I am reaching my health goals. |
| Help From Others | |
| Question 6 | I have someone whom I can call with questions about  my medicines. |
| Talking With Healthcare Team | |
| Question 7 | My doctor/nurse and I work together to make decisions. |
| Taking Medicines | |
| Question 8 | Have you taken a medicine more or less often than prescribed? |
| Question 9 | Have you skipped or stopped taking a medicine because you didn’t think it was working? |
| Question 10 | Have you skipped or stopped taking a medicine because it made you feel bad? |
| Question 11 | Have you skipped, stopped, not refilled, or taken less medicine because of the cost? |
| Question 12 | Have you not had medicine with you when it was time to take it? |

The English-translated version ^3^ is also provided for each item and response.

Questions 1 through 3 constitute the “Inconvenience/Forgetfulness” domain, questions 4 through 7 the “Treatment Beliefs” domain, and questions 8 through 12 the “Behavior” domain.

**Box S2. Japanese version of the Comprehensive Score for Financial Toxicity (COST)** ^1,2^

| Instruction sentence | Please circle one number per line to indicate your response as it applies |
| --- | --- |
| Question 1 | I know that I have enough money in savings, retirement, or assets to cover the costs of my treatment. |
| Question 2 | My out-of-pocket medical expenses are more than I thought they would be. |
| Question 3 | I worry about the financial problems I will have in the future as a result of my illness or treatment. |
| Question 4 | I feel I have no choice about the amount of money I spend on care. |
| Question 5 | I am frustrated that I cannot work or contribute as much as I usually do. |
| Question 6 | I am satisfied with my current financial situation. |
| Question 7 | I am able to meet my monthly expenses. |
| Question 8 | I feel financially stressed. |
| Question 9 | I am concerned about keeping my job and income, including work at home. |
| Question 10 | My illness or treatment has reduced my satisfaction with my present financial situation. |
| Question 11 | I feel in control of my financial situation. |
| Response options for questions | Not at all / A little bit / Somewhat / Quite a bit / Very much |

In this study, the term “cancer” was replaced with “illness” in question 10.
